# Modifiable Predictors of Sleep Quality in Multiple Sclerosis: A Prospective Cohort Study

**DOI:** 10.64898/2026.05.29.26354460

**Authors:** Marisa DelSignore, Shruthi Venkatesh, Wen Zhu, Matthew Goodman, Zongqi Xia

## Abstract

**Background:** Poor sleep quality is common in people with multiple sclerosis (pwMS) and reduces quality of life.

**Objectives:** To examine associations between modifiable factors and sleep quality in pwMS.

**Methods:** In a prospective clinic cohort (2017–2023), we evaluated whether baseline measures of disability, depression, fatigue, and pain were associated with poor sleep quality (Pittsburgh Sleep Quality Index, PSQI) cross-sectionally using covariate-adjusted linear regression, structural equation modeling (SEM), and LASSO logistic regression, and longitudinally using mixed-effects models.

**Results:** In this cohort (n=750; mean age 48.9 years; 80.3% women, 88.7% relapsing type), higher body mass index (β [95% CI]: 0.06 [0.01, 0.12], p=.001) and area deprivation index (6.78 [2.17, 11.39], p<.001) were associated with worse baseline PSQI scores. In adjusted analyses (n=730), disability, depression, fatigue, and pain were each associated with worse sleep. In SEM, pain had a moderate direct effect on sleep (β [95% CI]: 0.56 [0.48, 0.64], p<.001). LASSO models that included pain outperformed the benchmark (AUROC 0.741 vs 0.517). Longitudinally (n=382), time and higher baseline pain predicted worse sleep (β [95% CI]: time in months 0.04 [0.02, 0.06], p<.001; pain 0.36 [0.31, 0.41], p<.001).

**Conclusion:** Pain is a key, potentially modifiable driver of poor sleep quality in pwMS.

**Key Messages:**

◼ Sleep disturbance is common in people with MS and often worsens over time.
◼ Higher baseline pain severity was consistently associated with worse sleep quality, both cross-sectionally and longitudinally.
◼ Pain is a key modifiable driver of sleep disturbance and a potentially important therapeutic target for improving overall quality of life.

## INTRODUCTION

Sleep disturbance is prevalent among people with multiple sclerosis (pwMS), affecting 25-87% and diminishing quality of life.^1,2^ Common sleep disorders among pwMS include insomnia, sleep apnea, nocturnal movement disorders, narcolepsy, and nocturia.^3–9^ Sleep disturbance may result from inflammatory demyelination involving the sleep-wakefulness circuitry, elevated pro-inflammatory cytokines, adverse events due to disease-modifying therapies (DMTs) or symptomatic medications, MS-related symptoms (e.g., primary nocturia, leg spasms, muscle stiffness), disability progression, or co-existing primary sleep disorders and psychiatric illness.^10–14^

Poor sleep quality in MS is associated with disability, depression, fatigue, pain, and cognitive impairment.^2,15–20^ However, identifying and treating sleep problems remain challenging.^21^ Longitudinal data on sleep quality in pwMS are limited. Two preliminary studies demonstrated that sleep problems persisted over one year, though neither showed statistically significant changes over time.^22,23^ Evidence linking disease duration to poor sleep quality is also mixed.^17,24^

Given these gaps, we examined whether poor sleep quality correlates with other common MS symptoms (vs healthy controls), how baseline sleep quality relates to other symptoms using structural and prediction modeling, and what predicts longitudinal sleep quality trends.

## METHODS

### Participants

We included adult participants enrolled in a clinic cohort (Prospective Investigation of MS in the Three Rivers Region, PROMOTE; NCT02994121) between January 1, 2017, and November 17, 2023, including those with a neurologist-confirmed MS diagnosis and healthy controls. All participants had electronic health record (EHR) data linkage and completed at least one annual questionnaire capturing clinical history and patient-reported outcomes (PROs).

### Ethics Approval

The University of Pittsburgh Institutional Review Board approved this study (STUDY19080007). All participants provided written informed consent.

### Exposures

We assessed neurological function and disability, pain, depression, fatigue, and contentment at baseline and longitudinally using validated PROs that capture patient-reported symptoms and function and serve as proxies for the modeled domains. The index date was the first available sleep quality score (see below). We included other baseline PROs within 1 year of the index date. Instruments are described further in **S-Method 1**. The correlation structure among PROs in the MS cohort are shown (**S-Figure 1**).

#### Neurological function

Neurological function was quantified using the Patient Determined Disease Steps (PDDS),^25^ Multiple Sclerosis Rating Scale-Revised (MSRS-R),^26^ Patient Reported Outcomes Measurement Information System v1.2 Physical Function (PROMIS PF),^27^ and Functional Assessment of Multiple Sclerosis (FAMS) Mobility subscale.^28^

#### Mood

Mood was assessed using the Center for Epidemiologic Studies Depression Scale (CESD)^29^ and the FAMS Emotional Wellbeing subscale. ^28^

#### Fatigue

Fatigue was scored using the Modified Fatigue Impact Scale (MFIS-21),^30^ Perceived Deficits Questionnaire (PDQ),^31^ and the FAMS Thinking/Fatigue subscale.^28^

#### Pain

Pain was measured using the Medical Outcomes Study Pain Effects Scale (PES)^32^ and the FAMS Symptoms subscale.^28^

#### Contentment

Contentment was evaluated using the FAMS Contentment subscale.^28^

### Outcome

The primary outcome was sleep quality measured by the Pittsburgh Sleep Quality Index (PSQI), either as a continuous or dichotomous outcome (>5), with higher scores indicating worse sleep quality.^33^

### Covariates

To select *key* clinical or demographic covariates for downstream analyses, we performed univariate regression analyses using baseline PSQI score as the dependent outcome and each of the following potential predictors: age, sex, race, ethnicity, MS subtype, disease duration, DMT mechanism class and efficacy class, body mass index (BMI), comorbidities, and area deprivation index (ADI). We included covariate values within one year of the baseline PSQI where appropriate.

Participants represented the clinic population. We dichotomized race and ethnicity as non-Hispanic White and other minorities given the modest proportion of the latter. Neurologist-confirmed MS subtypes included relapsing-remitting MS (RRMS), clinically isolated syndrome (CIS), radiologically isolated syndrome (RIS), secondary progressive MS (SPMS), and primary progressive MS (PPMS). We grouped RRMS, CIS, and RIS into a non-progressive phenotype, and SPMS and PPMS into a progressive phenotype. Disease duration was calculated as years from first MS-related symptom onset to baseline. We categorized DMTs by mechanism and consensus efficacy class (high, standard, none) (**Table 1**). We calculated BMI from height and weight. We quantified comorbidity burden using the Elixhauser Comorbidity Index (ECI) based on ICD-9/10 codes documented in the EHR during the year before the earliest PSQI score (R package “comorbidity”).^34–36^ We measured socioeconomic disadvantage using ADI, a neighborhood-level measure of income, education, employment, and housing quality, derived from 2018–2022 American Community Survey data and participant zip codes using the Decentralized Geomarker Assessment for Multi-Site Studies (DeGAUSS) geocoder.^37^

**Table 1.** Study cohort characteristics.

|  | Multiple sclerosis (MS) | Control | P-value <sup>a</sup> |
| --- | --- | --- | --- |
| Number of patients (N) | 750 | 108 |  |
| Age at enrollment, mean ± SD | 48.86 ± 12.83 | 53.34 ± 14.29 | **0.001** |
| Male, N (%) | 148 (19.7) | 60 (55.6) | **<0.001** |
| Race/Ethnicity, N (%) |  |  |  |
| Non-Hispanic White | 699 (93.2) | 103 (95.4) | 1.00 |
| Race, N (%) |  |  | **0.04** |
| White | 706 (94.1) | 104 (96.3) |  |
| Black | 38 (5.1) | 1 (1.0) |  |
| Asian | 5 (0.7) | 3 (2.8) |  |
| Multi-racial | 1 (0.1) | 0 (0) |  |
| Ethnicity, N (%) |  |  | 0.61 |
| Hispanic or Latino | 5 (0.7) | 1 (1.0) |  |
| Not Hispanic or Latino | 742 (99.1) | 107 (99.1) |  |
| Not Sure / Not Reported | 1 (0.1) | 0 (0.0) |  |
| Enrollment Diagnosis, N (%) |  |  |  |
| *Non-Progressive MS* | 679 (90.5) | NA |  |
| Relapsing remitting MS | 665 (88.7) | NA |  |
| Clinically isolated syndrome | 9 (1.2) | NA |  |
| Radiologically isolated syndrome | 5 (0.7) | NA |  |
| *Progressive MS* | 68 (9.1) | NA |  |
| Primary progressive MS | 28 (3.7) | NA |  |
| Secondary progressive MS | 40 (5.3) | NA |  |
| Unknown subtype of MS | 3 (0.4) | NA |  |
| *Healthy Control* |  | 108 (100.0) |  |
| Disease-Modifying Therapy (DMT) mechanism <sup>b</sup>, N (%) |  |  |  |
| Alpha-4-integrin blocker | 133 (17.9) | NA |  |
| B cell depletion | 187 (25.2) | NA |  |
| Fumarate | 65 (8.8) | NA |  |
| Glatiramer acetate | 94 (12.7) | NA |  |
| Interferon-beta | 70 (9.4) | NA |  |
| Purine analog | 1 (0.1) | NA |  |
| Pyrimidine blocker | 9 (1.2) | NA |  |
| Sphingosine-1-phosphate receptor modulator | 14 (1.9) | NA |  |
| Other | 3 (0.4) | NA |  |
| None | 166 (22.4) | NA |  |
| DMT Efficacy Class <sup>c</sup> |  |  |  |
| High | 321 (43.3) | NA |  |
| Standard | 252 (34.0) | NA |  |
| None | 169 (22.8) | NA |  |
| Disease duration in years, mean ± SD | 15.96 ± 11.61 | NA |  |
| Body Mass Index, kg/m<sup>2</sup>, mean ± SD | 28.73 ± 7.72 | 28.33 ± 6.19 | 0.61 |
| Elixhauser Comorbidity Index, mean ± SD | 6.32 ± 3.56 | 5.71 ± 2.93 | 0.65 |
| Area deprivation index, mean ± SD | 0.29 ± 0.09 | 0.28 ± 0.08 | 0.51 |

a. P-values obtained from two-sample T tests.
b. The disease-modifying therapy (DMT) included the following under each mechanism: alpha4-integrin blocker (natalizumab), B cell depletion (ocrelizumab, ofatumumab, rituximab), fumarate (dimethyl fumarate, diroximel fumarate), glatiramer acetate, interferon-beta, purine analog (cladribine), pyrimidine blocker (teriflunomide), sphingosine-1-phosphate receptor modulator (fingolimod).
c. High-efficacy DMT included: alpha4-integrin blocker, B cell depletion, purine analog. Standard-efficacy DMT included: fumarate, glatiramer acetate, interferon-beta, sphingosine-1-phosphate receptor modulator, pyrimidine blocker.

### Statistical Analyses

#### Comparison of MS and Control

We compared pwMS and healthy controls using chi-square or Fisher’s exact tests for categorical variables and two-sample t-tests for continuous variables.

#### Cross-Sectional Analyses

We first used covariate-adjusted multiple linear regressions in the MS cohort to examine associations between PROs and baseline PSQI. We then fit a parsimonious structural equation model (**SEM**) to estimate directional paths among clinical domains (depression, sleep quality, fatigue, etc.).^38^ Directionality between domains was informed by clinical plausibility and prior literature and further evaluated by comparing alternative path specifications. We retained domains and paths that improved overall model fit, evaluated using chi-square, comparative fit index (CFI), Tucker-Lewis index (TLI), root mean square error of approximation (RMSEA), and standardized root mean square residual (SRMR).

We initially modeled each clinical domain as a latent construct using all available and relevant PROs for that domain (e.g., PDDS, MSRS-R, and PROMIS PF for disability; PES and FAMS Symptoms for pain). To reduce multicollinearity and improve model stability, we compared candidate indicators within each domain and selected a single validated measure that yielded the best overall model fit as the most parsimonious and clinically interpretable. The final model included PROMIS PF (disability), PES (pain), MFIS-21 (fatigue), CES-D (depression), FAMS Contentment (contentment), and PSQI (sleep). We evaluated multiple SEM specifications but reported only the final model, which had optimal performance.

Scores were z-standardized and aligned such that higher values reflected worse status where applicable. Models were estimated with maximum likelihood (lavaan).^39^

#### Machine Learning Prediction

We performed proportional sampling by outcome to establish training and held-out test datasets (70:30 ratio). Using least absolute shrinkage and selection operator (LASSO) logistic regression to predict poor baseline sleep quality as a binary outcome, we compared four models: (1) a benchmark model containing key covariates only, (2) key covariates plus the key predictors, (3) the key predictors only, and (4) all covariates plus the key predictors. We reported accuracy, precision, recall, F1 score, area under the receiver operating characteristic curve (AUROC), sensitivity, specificity, and false positive rate.

#### Longitudinal Analyses

We used linear mixed-effects models to examine associations between baseline predictors and repeated PSQI scores among pwMS who had at least two PSQI assessments. Fixed effects included baseline scores of key predictors, key covariates (selected by univariate regressions), months between PSQI assessments, and predictor-by-time interactions. We modeled intercepts and slopes as random effects to partially mitigate bias from variable follow-up. Because harmonized time-varying measures were not available cohort-wide, we modeled baseline pain both continuously with a pain-by-time interaction and categorically through stratified linear mixed-effects modeling by baseline pain severity group (none, mild, severe). In a subset with repeat pain measures, we descriptively compared paired trajectories of pain and sleep. For comparison, we also fit a model using dichotomized MS subtype (progressive vs non-progressive) as the primary predictor. Model selection was based on Akaike and Bayesian information criteria (AIC and BIC, respectively) relative to a covariates-only model. For sensitivity analysis, we used generalized estimating equations (GEEs), which provide population-averaged longitudinal estimates.

#### Statistical Software and Threshold

Analyses were performed in R (2023.06.1+524) or Python (3.11.4). Statistical significance was set at p<0.05 after Bonferroni correction for multiple comparisons.

### Code Availability

Analysis code is publicly available on GitHub.^40^

### Data Availability

The study data include linked EHR and registry data. Anonymous summary data are publicly available on GitHub.^40^ Sharing of de-identified EHR and registry data with qualified external researchers may be permissible with approval from the IRB and regulatory oversight agents of the healthcare system once an appropriate Data Use Agreements (DUA) between institutions is in place following reasonable request to the corresponding author.

## RESULTS

### Participant Characteristics

**Figure 1** provides an overview of the study, which included 750 pwMS and 108 healthy controls (**Table 1**). The MS group was younger (mean [SD]: pwMS, 48.86 [12.83]; controls, 53.34 [14.29]; p=.001) and had more female participants than controls (% female: pwMS, 80.3%; controls, 44.6%; p<.001). Groups were similar in race/ethnicity, BMI, comorbidity burden (ECI), and socioeconomic disadvantage (ADI). Most pwMS had a non-progressive subtype (n=679, 90.5%; predominantly RRMS, n=665, 88.7%) and many received high-efficacy DMT (n=321, 43.3%). Compared with controls, pwMS reported consistently worse PROs (**S-Table 1**): sleep quality (mean difference (d) [95% CI]: PSQI, d=1.58, [0.70, 2.46]), disability (PDDS, d=1.37 [1.06, 1.67]; MSRSR, d=4.82 [3.88, 5.76]; PROMIS PF, d=−8.55 [−11.05, −6.05]; FAMS Mobility, d=−5.40 [−6.53, −4.26]), depression (CESD, d=5.72 [3.47, 7.98]; FAMS Emotional Wellbeing, d=−2.60 [−3.54, −1.66]), contentment (FAMS Contentment, d=−1.97 [−3.26, −0.68]), fatigue (MFIS-21, d=14.13 [9.14, 19.12]; FAMS Thinking/Fatigue, d=−5.83 [−7.72, −3.94]; PDQ, d=2.58 [1.68, 3.48]), and pain (PES, d=3.69 [2.43, 4.94]; FAMS Symptoms, d=−2.22 [−3.35, −1.09]).

**Figure 1.**
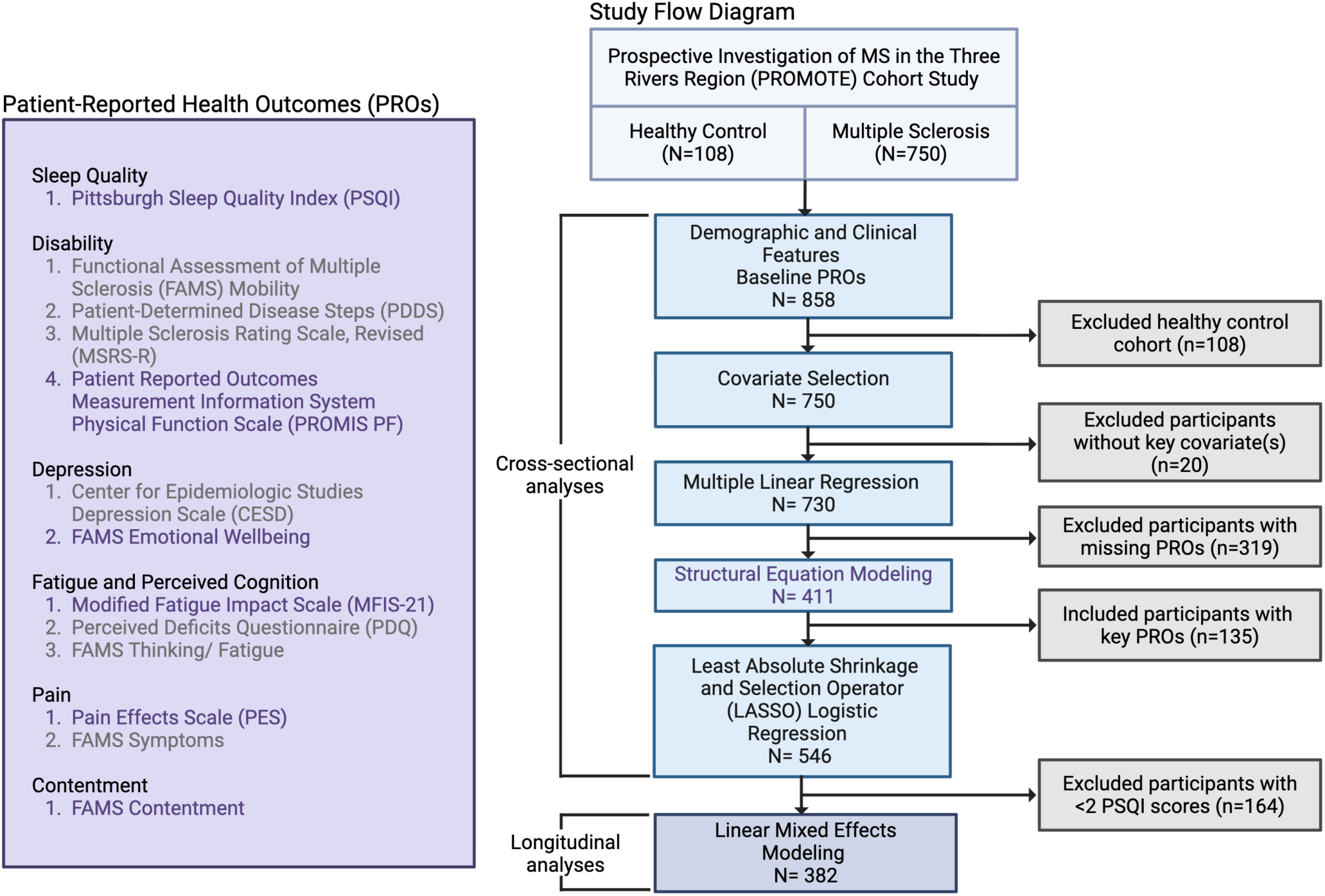
Study overview. Participants were selected from a clinic cohort that included people with multiple sclerosis and healthy controls. After excluding healthy controls and participants with missing data (e.g., key covariates), we performed cross-sectional analyses using multiple linear regression and structural equation modeling (SEM). SEM incorporated selected PROs (purple) to model latent constructs related to sleep quality. We used LASSO logistic regression to evaluate the performance of key predictor(s) on poor sleep quality. For longitudinal analysis, linear mixed-effects modeling examined the association between baseline predictors and repeated sleep quality scores. <u>Abbreviations</u>: *CESD*, Center for Epidemiologic Studies Depression; *FAMS*, Functional Assessment of Multiple Sclerosis; *LASSO*, Least Absolute Shrinkage and Selection Operator; *MFIS-21*, Modified Fatigue Impact Scale, 21 items; *MSRS-R*, Multiple Sclerosis Rating Scale, Revised; *PDDS*, Patient Determined Disease Steps; *PDQ*, Perceived Deficits Questionnaire; *PES*, Pain Effects Scale; *PROs*, Patient-Reported Outcomes; *PROMIS PF*, Patient Reported Outcomes Measurement Information System Physical Function; *PROMOTE*, Prospective Investigation of Multiple Sclerosis in the Three Rivers Region; *PSQI*, Pittsburgh Sleep Quality Index; *SEM*, Structural Equation Modeling.

### Selection of Key Covariates

Univariate regressions selected two key covariates associated with sleep quality in the MS cohort (n=750). Higher BMI and higher ADI were associated with worse baseline PSQI (regression coefficient β [95% CI]: BMI, 0.06 [0.01, 0.12], p=.001; ADI, 6.78 [2.17, 11.39], p<.001) (**S-Table 2**).

### MS Symptoms Associated with Baseline Poor Sleep Quality

In covariate-adjusted multiple linear regression analyses among pwMS (n=730), worse baseline sleep quality was consistently associated with greater disability (β [95% CI]: PDDS, 0.20 [-0.01, 0.41], p=.01; MSRS-R, 0.27 [0.19, 0.35], p<.001; PROMIS PF, −0.09 [−0.14, −0.05], p<.001; FAMS Mobility, −0.14 [−0.20, −0.08], p<.001), worse mood (CESD, 0.20 [0.16, 0.23], p<.001; FAMS Emotional Wellbeing, −0.26 [−0.32, −0.19], p<.001), worse fatigue (MFIS-21, 0.11 [0.09, 0.13], p<.001; PDQ, 0.37 [0.29, 0.45], p<.001; FAMS Thinking/Fatigue, −0.23 [−0.27, −0.18], p<.001), more severe pain (PES, 0.40 [0.33, 0.46], p<.001; FAMS Symptoms, −0.37 [−0.43, −0.30], p<.001), and lower contentment (FAMS Contentment, −0.26 [−0.32, −0.19], p<.001) (**S-Table 3**).

### MS Symptoms Driving Baseline Poor Sleep Quality

After optimizing the SEM (adequate fit across multiple parameters, **S-Table 4**), we report relations among key covariates, relevant PROs informing MS symptom severity, and baseline sleep quality in a MS subgroup with complete PROs (n=411) **(Figure 2, S-Table 5)**. Pain (PES) had the greatest direct effects on poor sleep quality (Standardized Direct Effect Estimate (θ) [95% CI]: 0.56 [0.48, 0.64], p<.001). Fatigue and pain each worsened depression (θ [95% CI]: 0.42 [0.33, 0.51], p<.001; 0.33 [0.24, 0.42], p<.001, respectively). Beyond its effect on sleep quality, pain also directly affected disability, fatigue and depression (θ [95% CI]: PROMIS PF, 0.55 [0.47, 0.63], p<.001; MFIS-21, 0.46 [0.37, 0.54], p<.001; FAMS Emotional Wellbeing: 0.33 [0.24, 0.42], p<.001). Neither key covariate significantly affected PSQI (θ [95% CI]: ADI, 0.05 [-0.05, 0.14], p=.32; BMI, 0.03 [-0.06, 0.11], p=.56).

**Figure 2.**
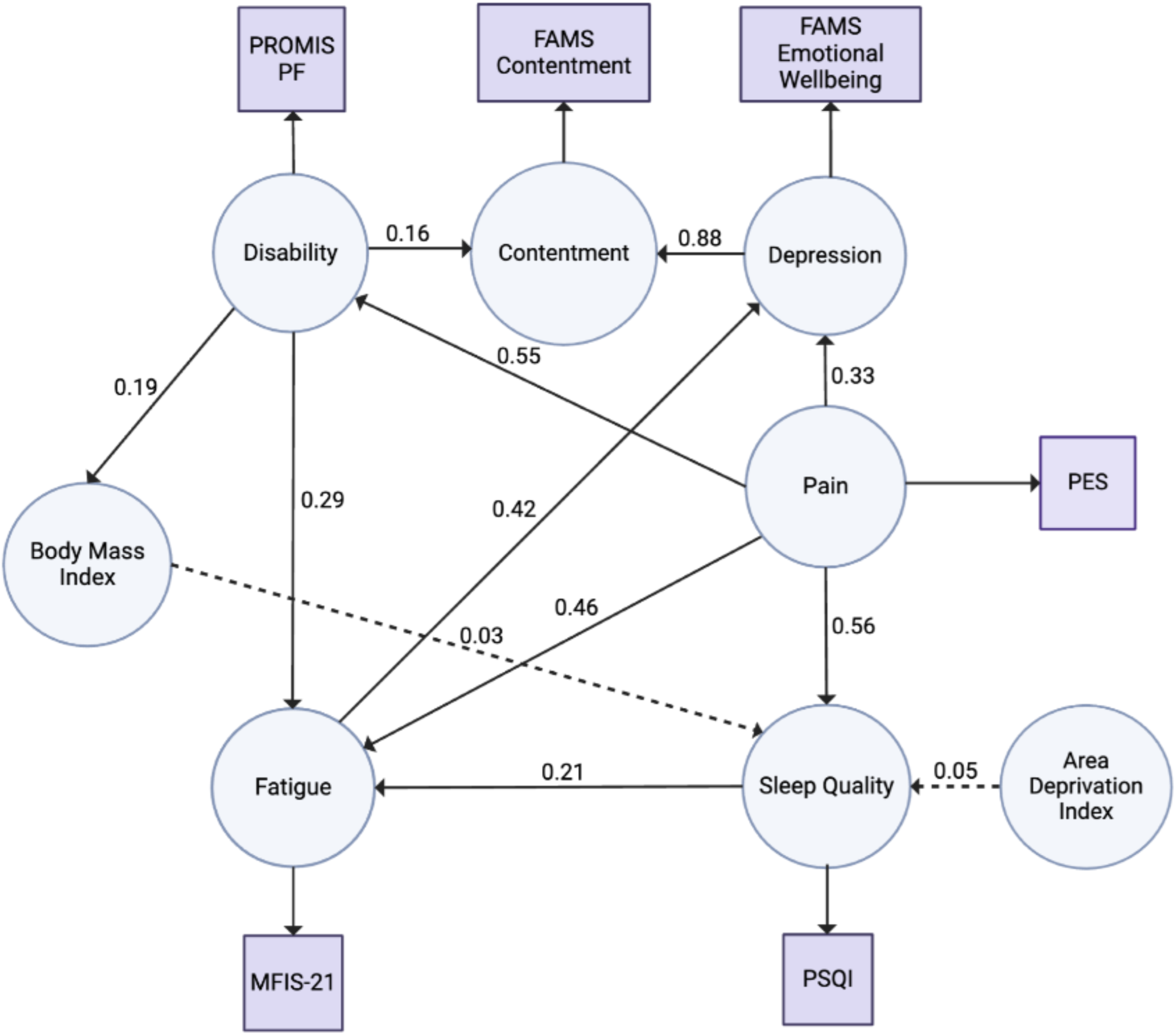
Directional relations among patient-reported outcomes and sleep quality (structural equation modeling of latent variables in the cross-sectional multiple sclerosis cohort). Using structural equation modeling, we examined relations among key clinical and demographic covariates, relevant patient-reported outcomes (PROs informing MS symptom severity), and sleep quality (**S-Table 5**). Domains (sleep quality, disability, fatigue, pain, depression, and contentment) were initially modeled as latent constructs using multiple related PROs. We then compared candidate indicators within each domain and selected a single PRO that yielded the best overall model fit. The final model illustrates directional associations between domains based on these selected indicators. Solid arrows indicate statistically significant associations, whereas dashed arrows indicate non-significant associations. Standardized direct effect estimates are shown. <u>Abbreviations</u>: *FAMS*, Functional Assessment of Multiple Sclerosis; *MFIS-21*, Modified Fatigue Impact Scale, 21 items; *PES*, Pain Effects Scale; *PROMIS PF*, Patient Reported Outcomes Measurement Information System Physical Function; *PSQI*, Pittsburgh Sleep Quality Index; *SEM*, Structural Equation Modeling.

### Prediction of Baseline Poor Sleep Quality

In the MS subgroup with available PROs selected by SEM (n=546 pwMS, 381 for training and 165 for test), we evaluated machine learning (LASSO) models to predict poor sleep quality (PSQI>5). Based on the SEM findings, we used baseline pain score (PES) as the primary predictor. The three models containing pain (pain plus all covariates, AUROC=0.755; pain plus BMI and ADI, AUROC=0.741; pain alone, AUROC=0.748) performed comparably and all outperformed the benchmark without pain (key covariates of BMI and ADI, AUROC=0.517) across performance indices **(Figure 3, S-Table 6)**. LASSO coefficients were consistent across models containing these variables (pain, 0.038–0.039; BMI, 0.003–0.006; ADI, 0) **(S-Table 7)**.

**Figure 3.**
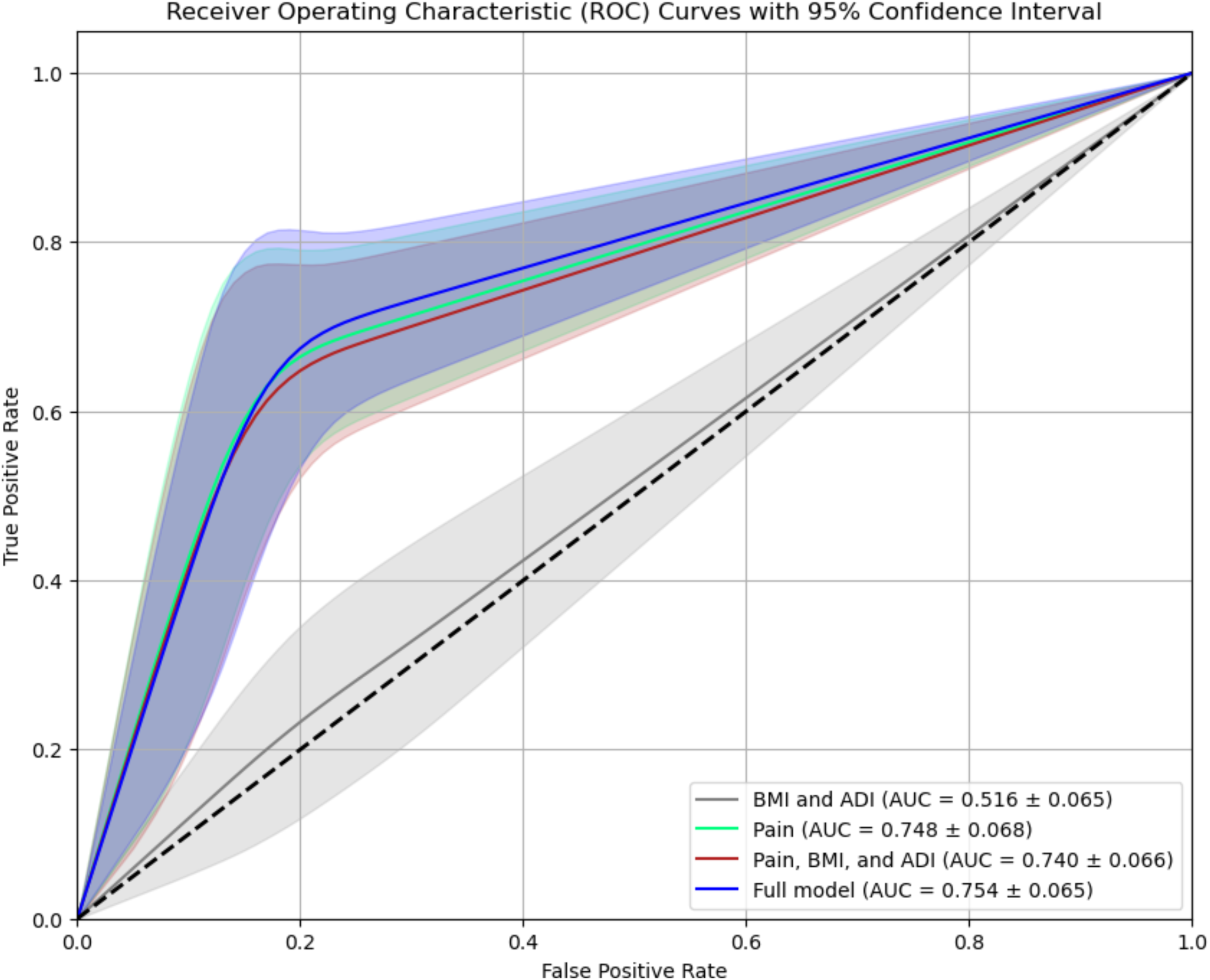
Machine learning model prediction of baseline poor sleep quality (in the cross-sectional multiple sclerosis cohort). Using a machine learning approach (least absolute shrinkage and selection operator, LASSO), we compared the performance of models containing pain versus the benchmark model without pain for predicting baseline poor sleep quality (PSQI>5). Pain was the primary predictor identified from structural equation modeling. First, we included all covariates (age at study enrollment, sex, race, ethnicity, body mass index [BMI], multiple sclerosis subtype at enrollment, disease-modifying therapy efficacy, disease duration, comorbidities, and area deprivation index [ADI]) and pain (Model 1: Full Model; blue curve). Second, we included key covariates identified through univariate regressions (BMI and ADI) and pain (Model 2: Pain, BMI, and ADI; red curve). Third, we included pain only (Model 3: Pain; green curve). Finally, we included key covariates (BMI and ADI) without pain (Model 4: BMI and ADI; black curve). In receiver operating characteristic (ROC) analysis, the area under the curve among Models 1, 2, and 3 were comparable, with all three outperforming Model 4.

### Longitudinal Changes in Sleep Quality

In an MS subgroup with ≥2 PSQI assessments (n=382), we assessed the association between baseline pain and longitudinal changes in sleep quality using a linear mixed-effects model to account for patient-level variability, adjusting for key covariates (**Table 2**). Higher baseline pain was associated with worse sleep quality (β [95% CI]: 0.36 [0.31, 0.41], p<.001). Based on the effect of time (months) (β [95% CI]: 0.04 [0.02, 0.06], p<.001), PSQI scores increased (i.e., worsened) by 0.04 per month among pwMS with a baseline pain (PES) score of 0. Based on the negative interaction term between time elapsed from baseline and baseline pain (β [95% CI]: −0.004 [−0.005, −0.002], p<.001), the monthly rate of change (i.e., slope) decreases by 0.004 for each 1-unit increase in baseline pain. Thus, higher baseline pain is associated with a smaller longitudinal change in sleep quality. Put another way, the cross-sectional association between baseline pain and sleep quality shrinks by 0.004 for each additional month of follow-up such that the gap between pwMS having higher- versus lower baseline pain narrows over time. The interpretation requires caution (see **Discussion**).

**Table 2.** Covariate-adjusted linear mixed-effects model examining the association between baseline pain (continuous Pain Effects Scale score) and longitudinal changes in sleep quality (in the longitudinal multiple sclerosis cohort).

| Measure | Regression Coefficient | 95% CI | P-value |
| --- | --- | --- | --- |
| Time elapsed from baseline (months) | 0.04 | [0.02, 0.06] | **<0.001** |
| Pain (baseline) | 0.36 | [0.31, 0.41] | **<0.001** |
| Area deprivation index (baseline) | 2.36 | [-1.05, 5.76] | 0.17 |
| Body mass index (baseline), kg/m<sup>2</sup> | 0.03 | [-0.01, 0.07] | 0.18 |
| Time elapsed from baseline X pain (baseline) interaction | -0.004 | [-0.005, -0.002] | **<0.001** |

To assess longitudinal changes in sleep quality (Pittsburgh Sleep Quality Index, PSQI), we used *fixed* effects to model time elapsed from baseline, pain severity (Pain Effects Scale, PES), area deprivation index, body mass index, and the interaction term, and *random* effects to model the intercept and slope of the time interval variable. Baseline was defined as the first PSQI score.

Sensitivity analyses showed consistent findings. When stratifying pwMS by baseline pain, longitudinal sleep quality (PSQI) changes differed. Sleep quality worsened over time in the no-pain group (β [95% CI]: 0.02 [0.001, 0.03], p=.03), while it did not change significantly in the mild-pain group (β [95% CI]: −0.02 [−0.04, 0.002], p=.08) and improved slightly in the severe-pain group (β [95% CI]: −0.06 [−0.08, −0.03], p<.001) (**S-Table 8**). Compared to linear mixed-effects models not containing baseline pain (i.e., key covariates only, or MS subtype plus key covariates), the main covariate-adjusted linear mixed-effects model with baseline pain (**Table 2**) achieved better fit based on AIC and BIC (**S-Table 9**). When modeling longitudinal sleep quality changes using the less restrictive generalized estimating equations, the findings largely replicated the linear mixed-effects models (**S-Tables 9–10**).

In exploratory analyses of pwMS with repeat sleep quality and pain assessments, sleep quality worsened over time in those who started without pain, while it remained largely stable in the baseline mild and severe pain groups. Pain remained mostly stable over time (**S-Figure 2**).

## DISCUSSION

In this prospective clinic cohort study, pain was a primary driver of poor sleep quality in pwMS. Higher baseline pain severity was consistently associated with poor sleep quality cross-sectionally and longitudinally. Sleep quality in patients with lower baseline pain worsened over time, suggesting progressive disruption that may reflect neuroinflammatory, disability-related, or psychosocial factors.

Applying SEM to validated PROs, this study moved beyond symptom clustering to model directional relations among common MS symptoms.^41^ Although prior cluster analyses linked neurological impairment, fatigue, pain, depression, and sleep disturbances, this study identified the strongest predictor of poor sleep quality.^18,42–44^ In this framework, pain emerged as the primary predictor, consistent with the prior literature and supported by biological plausibility and the clinical overlap between sleep disruption and MS-related pain syndromes such as trigeminal neuralgia, neuropathic pain, Lhermitte’s phenomenon, tonic spasms, back pain, and headache.^5,45,46^

In our longitudinal analysis, pwMS with mild or severe baseline pain had poor sleep quality that largely persisted over time, while those without baseline pain showed a gradual worsening in sleep quality. The prior literature suggests that pain may have a stronger correlation with poor sleep later in the MS course.^24^ Our design does *not* establish causality, and the pain-by-time interaction has potential alternative interpretations, including regression to the mean, unmeasured time-varying confounders (e.g., pwMS with more severe pain are more likely to receive pain- or sleep-targeting interventions), and informative attrition (e.g., pwMS with the worst pain severity and sleep quality may be lost to follow-up). Moreover, factors such as relapse activity and disability progression may influence both pain and sleep. Thus, our longitudinal findings are best viewed as hypothesis-generating. Future studies incorporating time-varying clinical covariates, treatment exposures, and additional modeling strategies (e.g., non-linear time, splines) may clarify these complex relationships.

High ADI and BMI were key covariates associated with poor sleep quality in MS. These findings are consistent with prior work linking socioeconomic disadvantage to poor sleep quality in the general population, possibly more strongly in pwMS.^47,48^ Evidence linking obesity and poor sleep in MS has been mixed, though obesity is a known risk factor for obstructive sleep apnea and nocturnal hypoxemia^2,49–51^ as well as MS disability. In contrast, sleep quality was not associated with sex or DMT in this study, consistent with prior reports.^15,17,52^

Sleep problems in MS are often under-recognized and suboptimally treated despite their impact on quality of life. Our study calls particular attention to patients with moderate or severe pain, who are at heightened risk for poor sleep quality. Although many pwMS screen positive for obstructive sleep apnea, insomnia, or restless legs syndrome, these conditions often remain undiagnosed.^17,21^ Moreover, sleep-directed medications may have limited benefits without diagnosis-guided care.^53^ Comprehensive MS care should include routine screening and formal diagnosis of common sleep conditions. MS specialists should collaborate with sleep specialists to deliver mechanism-based management with emphasis on nonpharmacologic strategies (e.g., sleep hygiene coaching, cognitive behavioral therapy), parallel management of exacerbating factors through a multidisciplinary approach (e.g., adequate management of pain and nocturia), and judicious use of sleep medications.^54–56^

Our study has limitations. First, although we used multiple complementary modeling approaches and conservative inference to improve robustness, findings from this single center cohort require external validation in larger multicenter cohorts. Second, the dataset lacked information on anxiety levels or symptomatic MS therapies (e.g., methylprednisolone, baclofen, or gabapentin) that may additionally influence sleep quality.^11,57^

Third, sleep quality based on a validated PRO instrument and triangulated with other related PROs captures important patient experience, but discordance with objective measures could bias findings.^16,58^ Future studies could assess sleep quality using polysomnography or digital phenotyping with passive sensors such as actigraphy or smartphones. Fourth, our parsimonious SEM approach used single-indicator domains to reduce multicollinearity. Future studies could explore more complex multi-indicator models. Finally, to avoid overfitting, longitudinal analyses modeled baseline predictors of sleep quality trajectories given insufficient longitudinal covariates (e.g., post-baseline pain assessments, pain or sleep treatment, relapse, or disability progression). We partially addressed this limitation by fitting linear mixed-effects models with baseline pain as a continuous predictor (including a pain-by-time interaction), conducting stratified analyses by baseline pain severity category, and performing an exploratory analysis of paired pain and sleep trajectories. Future work should account for time-varying covariates within a causal framework.

## CONCLUSION

Our study identifies pain as a key modulator of poor sleep quality and a potentially treatable target to improve quality of life for pwMS. Future randomized clinical trials could confirm this hypothesis by comparing more intensive pain management with routine care on sleep quality and overall quality of life in pwMS.

## Data Availability

The study data include linked EHR and registry data. Anonymous summary data are publicly available on GitHub. Sharing of de-identified EHR and registry data with qualified external researchers may be permissible with approval from the IRB and regulatory oversight agents of the healthcare system once an appropriate Data Use Agreements (DUA) between institutions is in place following reasonable request to the corresponding author.

## ACKNOWLEDGEMENTS

We thank Drs. Judy Chang, Yanshan Wang, and Scott Rothenberger for their feedback and methodological guidance during the development of this work.

## SUPPLEMENTARY METHODS

**S-Method 1.** Patient-reported outcome measures.

### Neurological function

Patient Determined Disease Steps (**PDDS**): The PDDS is a single-item instrument that measures gait impairment on an ordinal scale ranging from normal (0) to bedridden (8). Developed by the North American Research Committee on MS (NARCOMS) registry to measure patient-reported disability accumulation in pwMS and derived from Disease Steps,^58^ the PDDS correlates with and complements the clinician-rated Expanded Disability Status Scale (EDSS).^24^

Multiple Sclerosis Rating Scale-Revised (**MSRS-R**): The MSRS-R assesses the global burden of neurological symptoms across eight domains: walking, upper limb function, vision, speech, swallowing, cognition, sensory, bladder, and bowel function.^25^ Each symptom domain is scored on a scale from 0 to 4, with the sum of domain subscores ranging from 0 to 32. Higher domain subscores and global scores indicate greater neurological symptom burden and disability.

Patient Reported Outcomes Measurement Information System Physical Function v1.2 (**PROMIS PF**): PROMIS PF is a generalizable measure of physical function across health and diseases.^26,59^ It measures upper extremity function (dexterity), lower extremity function (mobility), and central region function (neck, back) as well as the ability to perform complex activities spanning multiple physical domains (i.e., activities of daily living). PROMIS PF is reported as a T-score ranging from 0 to 100, where a score of 50 is the average for the general United States population. Higher T-scores indicate *better* physical function.

### Mood

Center for Epidemiologic Studies Depression Scale (**CESD**): CESD is a 20-item scale that measures depressive symptoms such as crying, loneliness, and low self-esteem, each rated on a 4-point scale (0–3), with total scores ranging from 0 to 60. Higher total scores indicate greater depressive symptom severity, and scores of 16 or above suggest clinically relevant depression.^28^

### Fatigue

Fatigue is a broad construct with both physical and psychological components. The MS Council for Clinical Practice Guidelines defines fatigue as: a “subjective lack of physical and/or mental energy that is perceived by the individual or caregiver to interfere with usual and desired activities.”^60^

Modified Fatigue Impact Scale (**MFIS-21**): The MFIS-21 is a 21-item assessment of fatigue across physical, cognitive, and psychosocial domains. Each item uses a 5-point Likert scale (0–4), with total scores ranging from 0 to 84. The modified version was adapted from the Fatigue Impact Scale when it was incorporated into the MS Quality of Life Inventory (MSQLI).^29,31,61^ A higher MFIS-21 score indicates a greater impact of fatigue on quality of life.

Perceived Deficits Questionnaire (**PDQ**): The PDQ is a 20-item scale that assesses subjective cognitive functioning in pwMS through four subscales: attention, retrospective memory, prospective memory, and planning and organization.^22^ Each item is scored on a 5-point scale (0–4), with higher total scores (0–80) indicating greater perceived cognitive impairment. Prior studies have reported that perceived cognitive performance and PDQ scores correlate more strongly with measures of depression and fatigue in pwMS than with measures of objective cognitive performance.^62,63^

### Pain

Pain Effects Scale (**PES**): The PES is a 6-item scale that assesses the effects of pain on daily functioning in pwMS. Each item is scored on a 5-point scale (1–5), with higher total scores (6–30) indicating worse pain severity. We also used PES scores to classify pain groups: no pain (6), mild pain(7–18), and severe pain (19–30).

### Quality of Life

Functional Assessment of Multiple Sclerosis (**FAMS**): The FAMS is a 59-item comprehensive quality-of-life measure that assesses seven subscales: mobility, symptoms, emotional well-being (depression and mood), contentment, thinking/fatigue, family/social well-being, and additional concerns. The symptoms subscale includes three specific pain items and a headache item. Items are rated on a 5-point Likert scale (from 0 = “not at all” to 4 = “very much”). Higher scores indicate better quality of life.

### Sleep Quality

Pittsburgh Sleep Quality Index (**PSQI**): The PSQI is a 19-item measure of subjective sleep quality and disturbances across seven components: sleep quality, latency, duration, habitual sleep efficiency, disturbances, use of sleep medication, and daytime dysfunction.^32,64^ Items are rated on a 4-point scale (0–3), with higher total score (0–21) indicating worse sleep quality. We quantified sleep quality using the PSQI as both a continuous outcome and a dichotomous outcome where total scores >5 indicate moderate-severe poor sleep quality.

## SUPPLEMENTARY FIGURES

**S-Figure 1.**
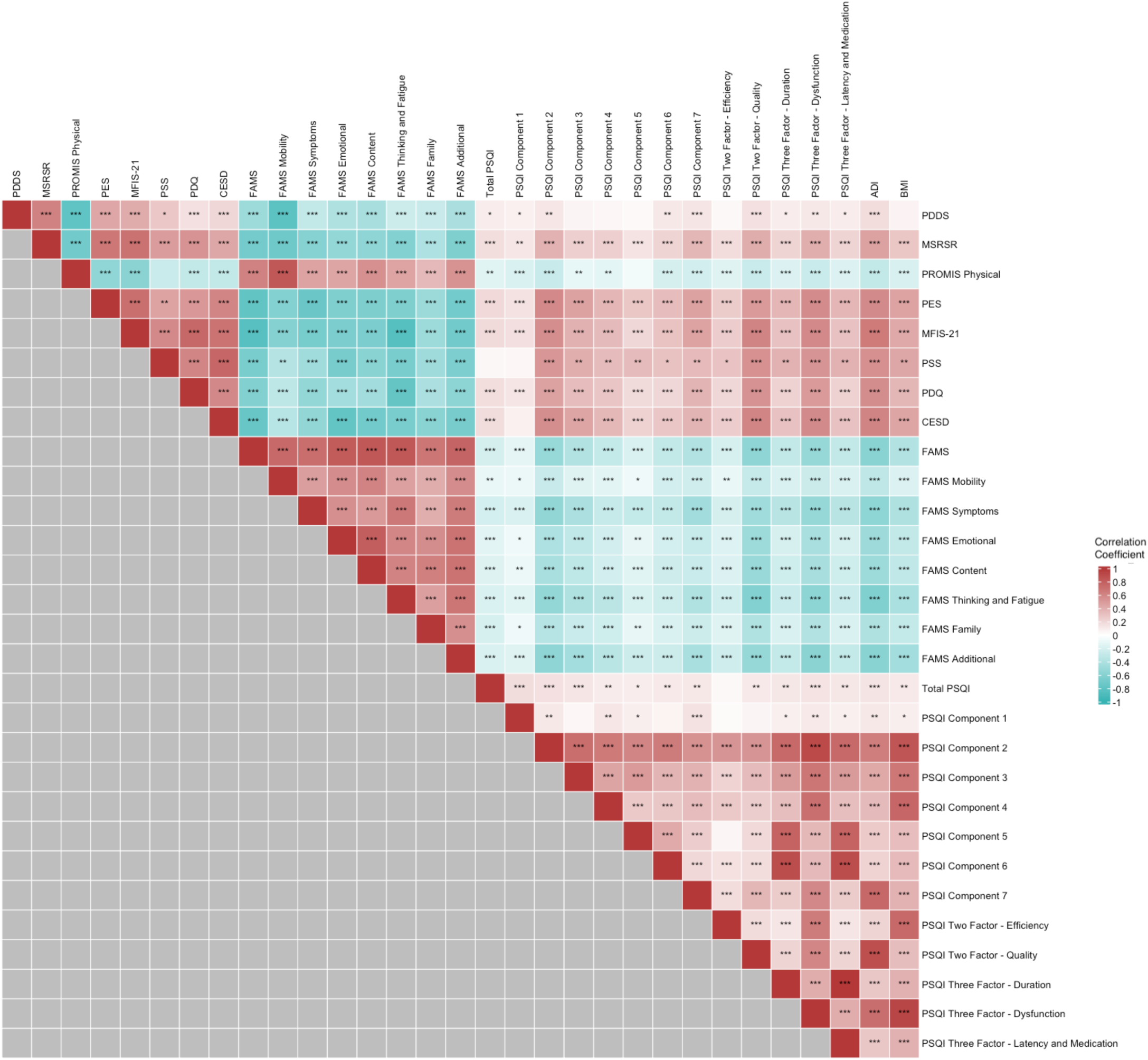
Correlation structure of patient-reported outcomes and key covariates in the multiple sclerosis cohort. P-value significance threshold: * p < 0.05, ** p < 0.01, *** p < 0.001. <u>Abbreviations</u>: *ADI*, Area Deprivation Index; *BMI*, Body Mass Index; *CESD*, Center for Epidemiologic Studies Depression; *FAMS*, Functional Assessment of Multiple Sclerosis; *LASSO*, Least Absolute Shrinkage and Selection Operator; *MFIS-21*, Modified Fatigue Impact Scale, 21 items; *MSRS-R*, Multiple Sclerosis Rating Scale, Revised; *PDDS*, Patient Determined Disease Steps; *PDQ*, Perceived Deficits Questionnaire; *PES*, Pain Effects Scale; *PROs*, Patient-Reported Outcomes; *PROMIS PF*, Patient Reported Outcomes Measurement Information System Physical Function; *PSQI*, Pittsburgh Sleep Quality Index.

**S-Figure 2.**
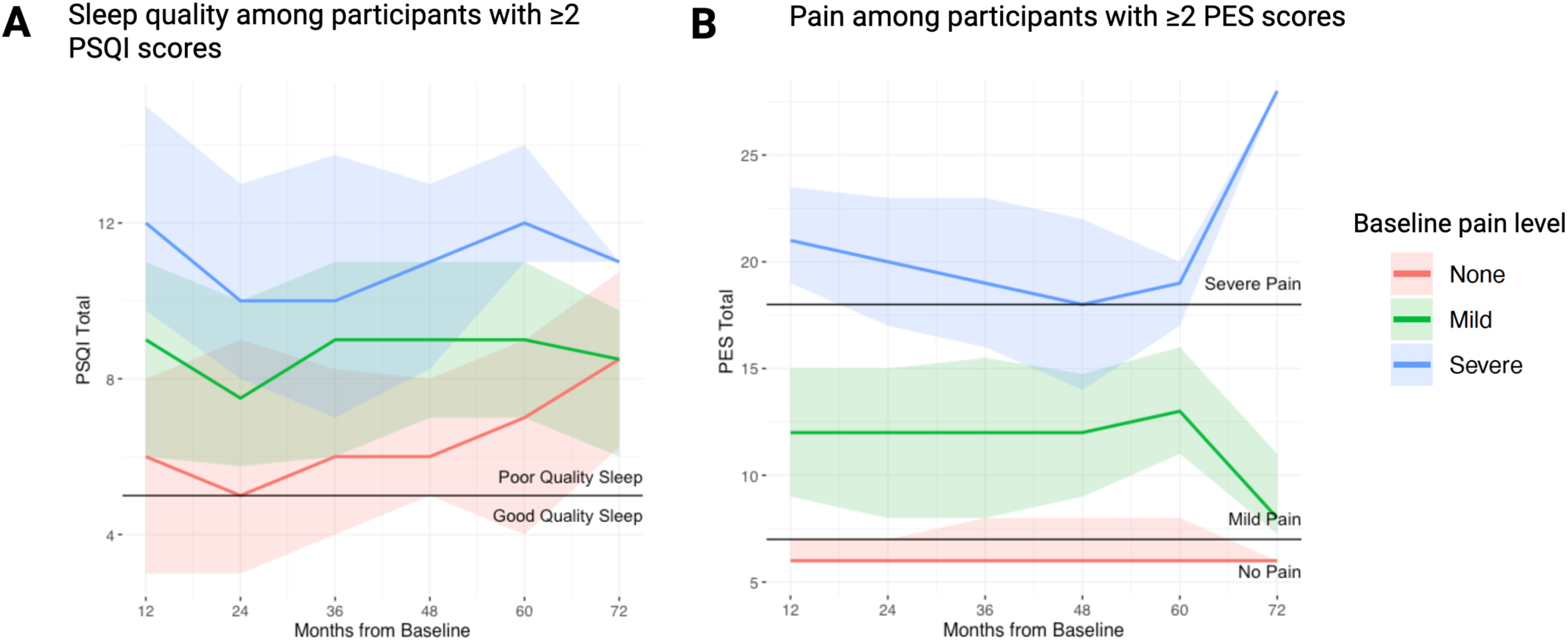
Longitudinal trends of sleep quality and pain stratified by baseline pain severity. We examined longitudinal trends in sleep quality and pain by baseline pain severity in the MS cohort. Sleep quality was measured using the Pittsburgh Sleep Quality Index (PSQI), and pain was measured using the Pain Effects Scale (PES). Baseline was defined as each participant’s first available assessment for the corresponding measure. Time from baseline was grouped into yearly intervals and plotted at the interval endpoint; for example, 12 months represents assessments from 0–12 months after baseline, and 24 months represents assessments from >12–24 months after baseline. Baseline pain was categorized as none (PES 0–6), mild (PES 7–18), or severe (PES 19–30). Lines represent median scores, and shaded regions represent the interquartile range. Horizontal reference lines indicate the PSQI threshold for poor sleep quality and PES thresholds for mild and severe pain. Panel A shows longitudinal PSQI scores among participants with at least two PSQI scores during the 72-month observation window (n=392). Panel B shows longitudinal PES scores among participants with at least two PES scores during the 72-month observation window (n=371). Because follow-up duration varied across participants, later intervals include fewer observations and should be interpreted descriptively.

## SUPPLEMENTARY TABLES

**S-Table 1.**
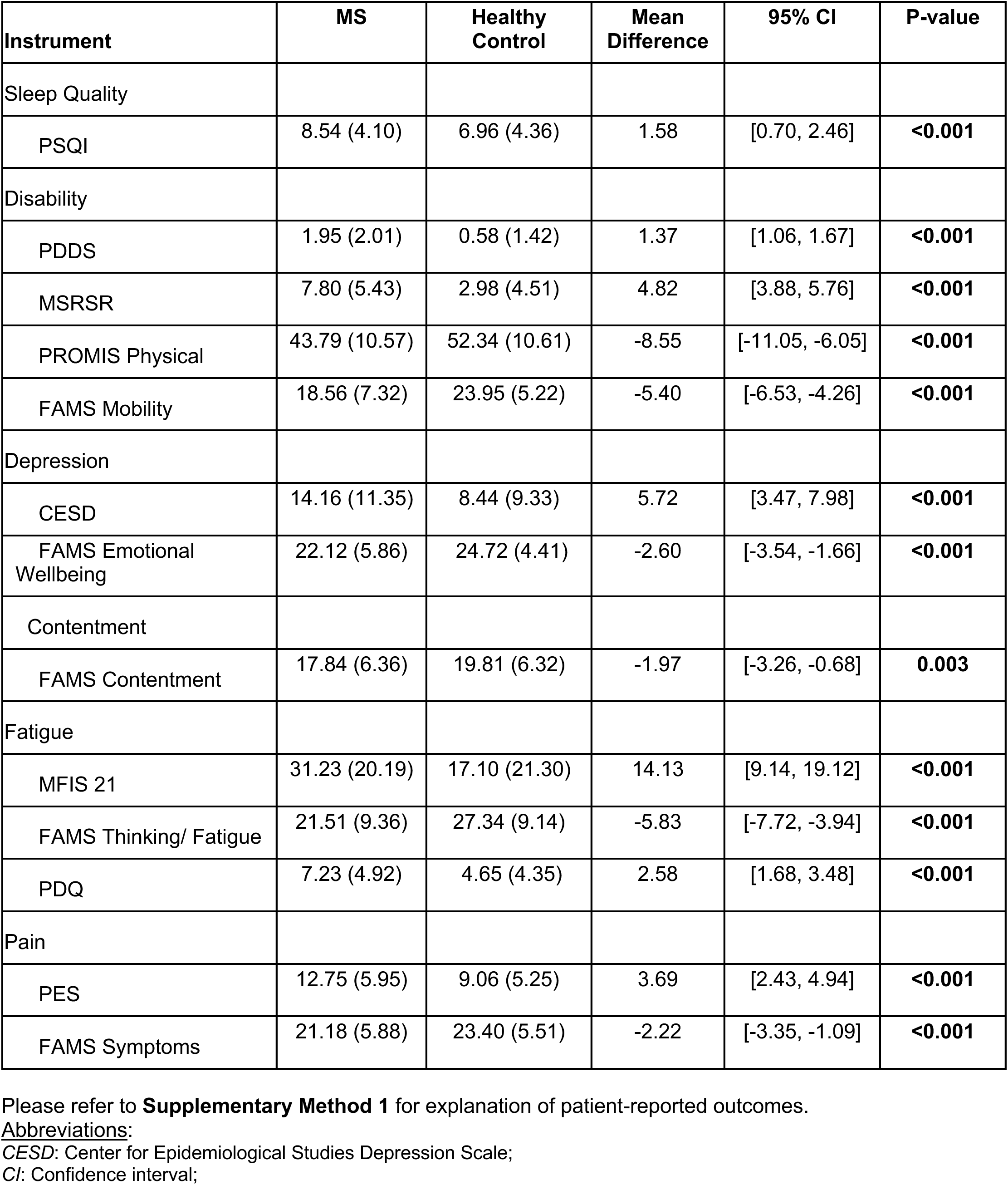
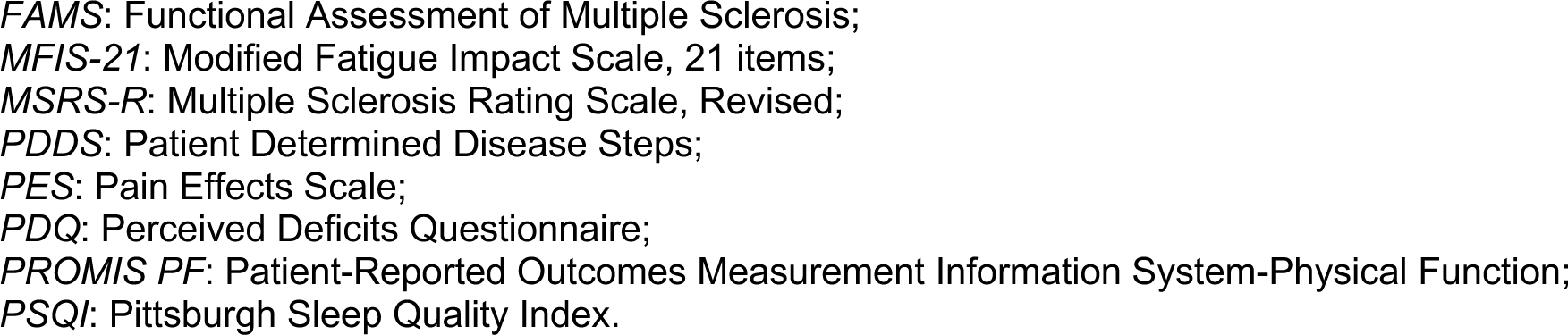
Baseline patient-reported outcomes in MS and healthy control groups (Welch two-sample t-test).

**S-Table 2.**
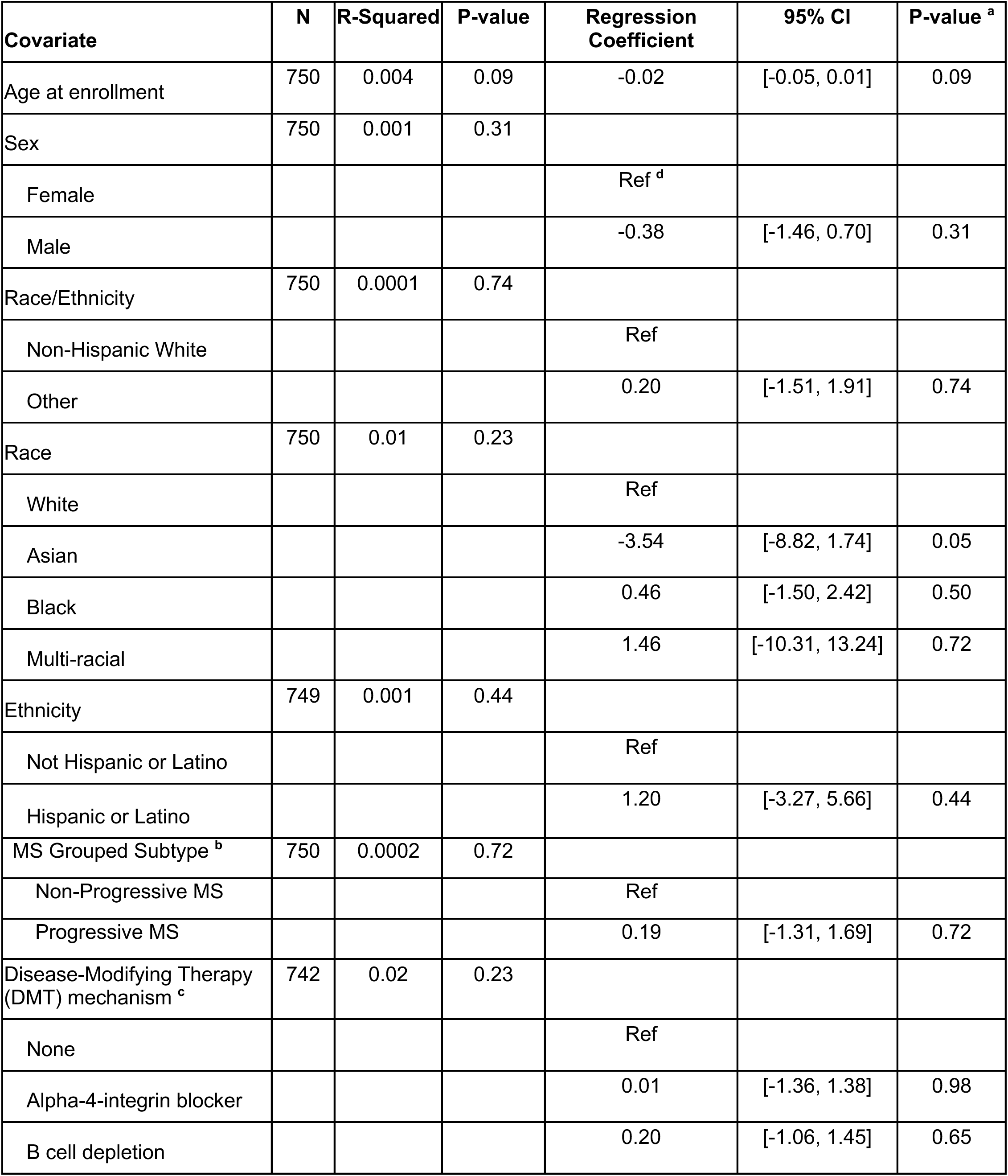
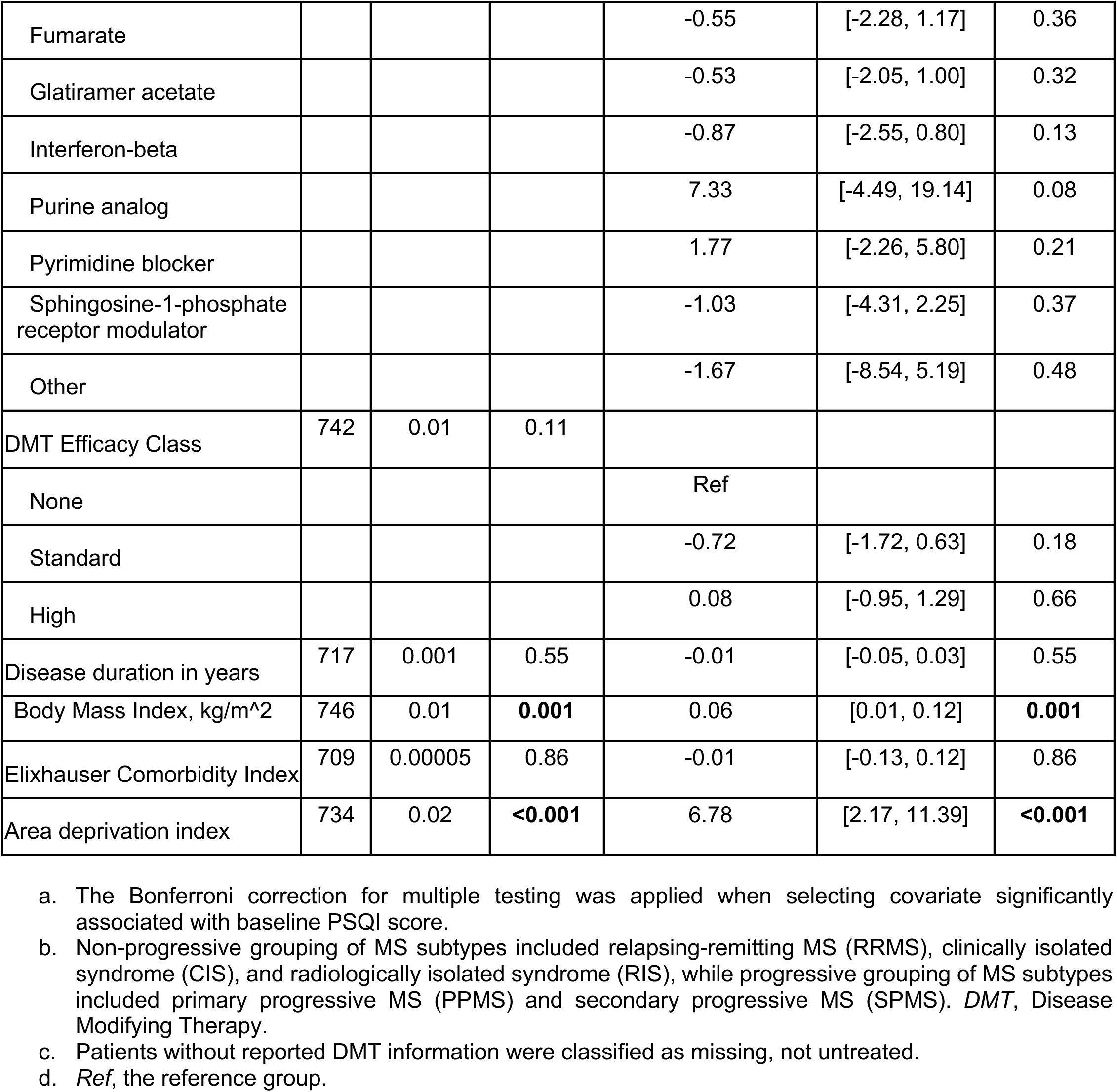
Selection of key clinical or demographic covariates associated with baseline PSQI scores in the multiple sclerosis group (univariate linear regression).

**S-Table 3.** Multiple sclerosis symptoms (patient-reported outcome scores) associated with baseline sleep quality (PSQI) scores (covariate-adjusted linear regression).

| **Measure <sup>a</sup>** | **N** | **Regression Coefficient <sup>b</sup>** | **95% CI** | **P-value <sup>c</sup>** |
| --- | --- | --- | --- | --- |
| Disability |  |  |  |  |
| PDDS | 728 | 0.20 | [-0.01, 0.41] | **0.01** |
| MSRS-R | 730 | 0.27 | [0.19, 0.35] | **<0.001** |
| PROMIS PF | 501 | -0.09 | [-0.14, -0.05] | **<0.001** |
| FAMS Mobility | 697 | -0.14 | [-0.20, -0.08] | **<0.001** |
| Depression |  |  |  |  |
| CESD | 603 | 0.20 | [0.16, 0.23] | **<0.001** |
| FAMS Emotional Wellbeing | 730 | -0.26 | [-0.33, -0.20] | **<0.001** |
| Contentment |  |  |  |  |
| FAMS Contentment | 730 | -0.26 | [-0.32, -0.19] | **<0.001** |
| Fatigue |  |  |  |  |
| MFIS-21 | 603 | 0.11 | [0.09, 0.13] | **<0.001** |
| FAMS Thinking/ Fatigue | 697 | -0.23 | [-0.27, -0.18] | **<0.001** |
| PDQ | 730 | 0.37 | [0.29, 0.45] | **<0.001** |
| Pain |  |  |  |  |
| PES | 603 | 0.40 | [0.33, 0.46] | **<0.001** |
| FAMS Symptoms | 730 | -0.37 | [-0.43, -0.30] | **<0.001** |

- Please refer to **Supplementary Method 1** for explanation of patient-reported outcomes. - All tests were adjusted for key covariates (area deprivation index and body mass index). - The Bonferroni correction for multiple testing was applied for determining statistical significance.

**S-Table 4.**
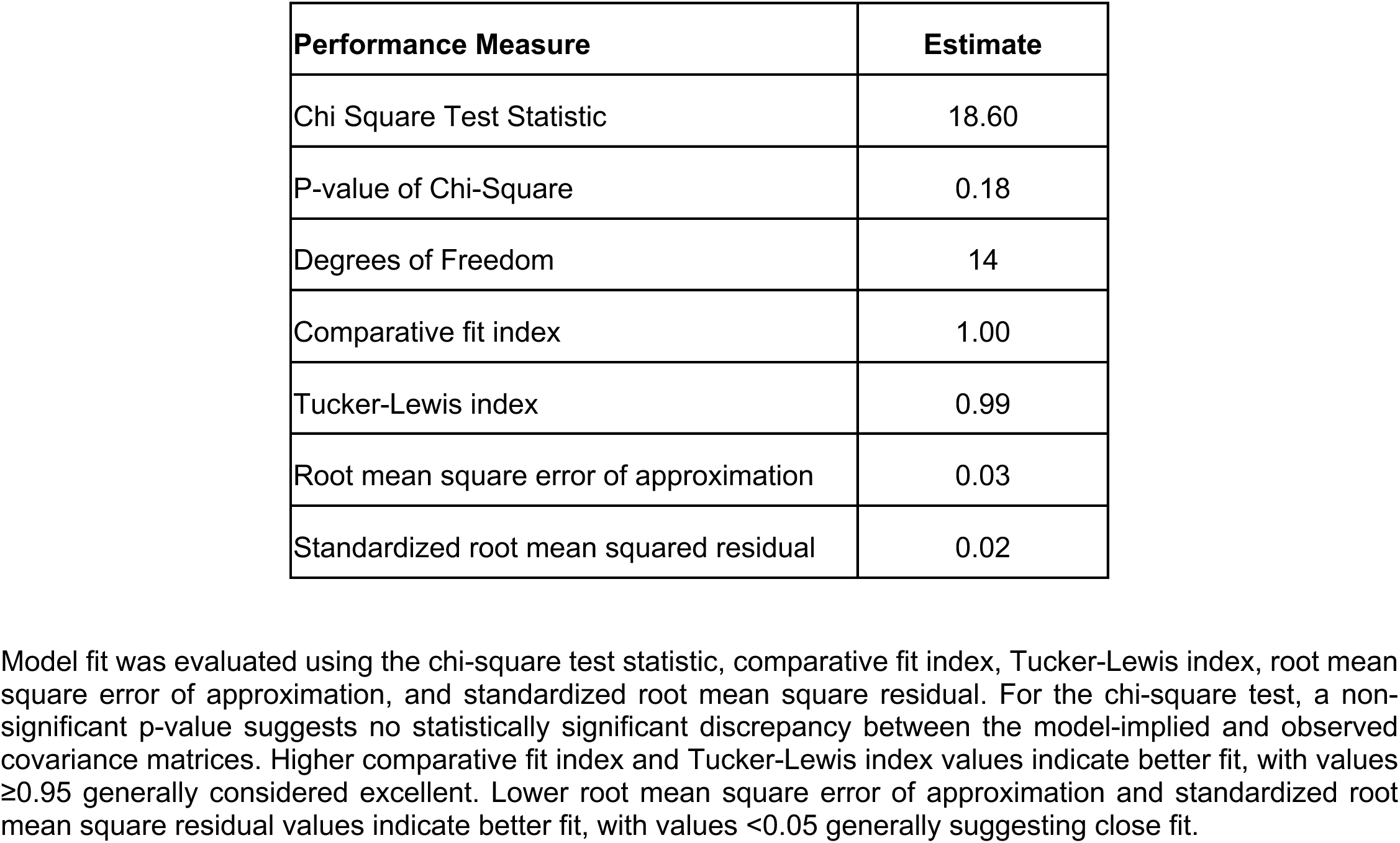
Performance indices of the final multiple sclerosis symptoms structural equation model.

| Performance Measure | Estimate |
| --- | --- |
| Chi Square Test Statistic | 18.60 |
| P-value of Chi-Square | 0.18 |
| Degrees of Freedom | 14 |
| Comparative fit index | 1.00 |
| Tucker-Lewis index | 0.99 |
| Root mean square error of approximation | 0.03 |
| Standardized root mean squared residual | 0.02 |

Model fit was evaluated using the chi-square test statistic, comparative fit index, Tucker-Lewis index, root mean square error of approximation, and standardized root mean square residual. For the chi-square test, a non-significant p-value suggests no statistically significant discrepancy between the model-implied and observed covariance matrices. Higher comparative fit index and Tucker-Lewis index values indicate better fit, with values $\geq 0.95$ generally considered excellent. Lower root mean square error of approximation and standardized root mean square residual values indicate better fit, with values $< 0.05$ generally suggesting close fit.

**S-Table 5.**
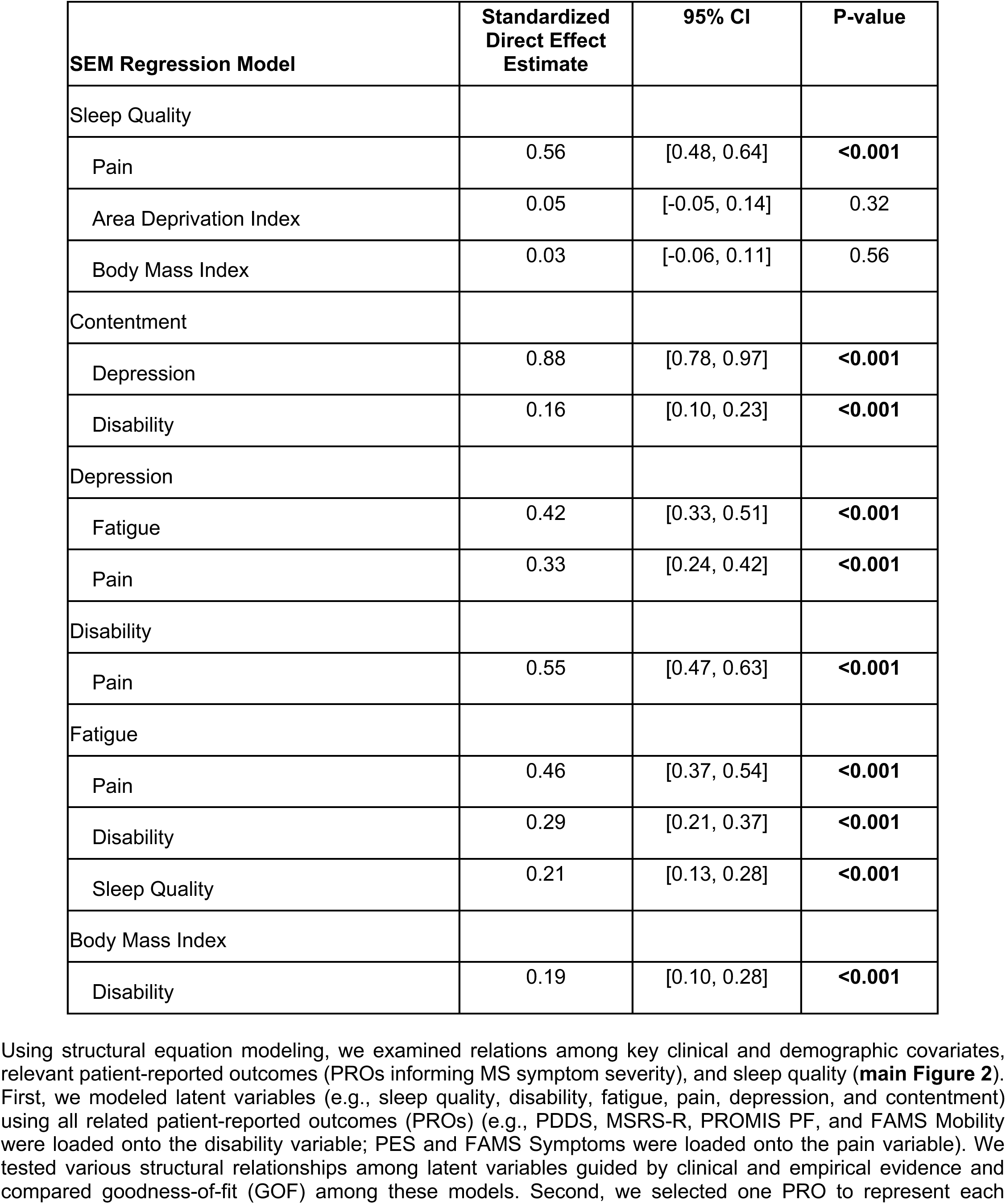
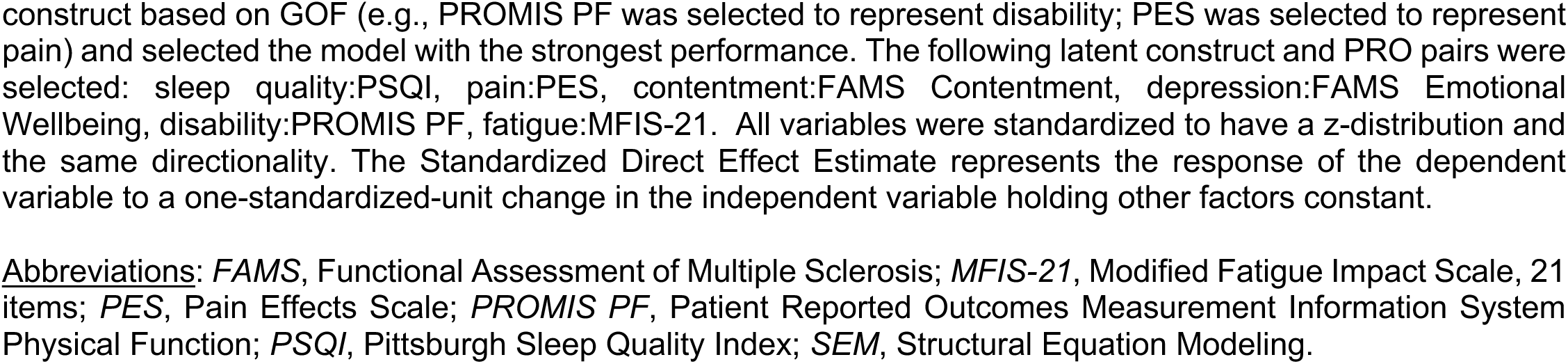
Structural equation model (SEM) of loaded multiple sclerosis patient-reported outcome (PRO) indicator as latent construct.

**S-Table 6.**
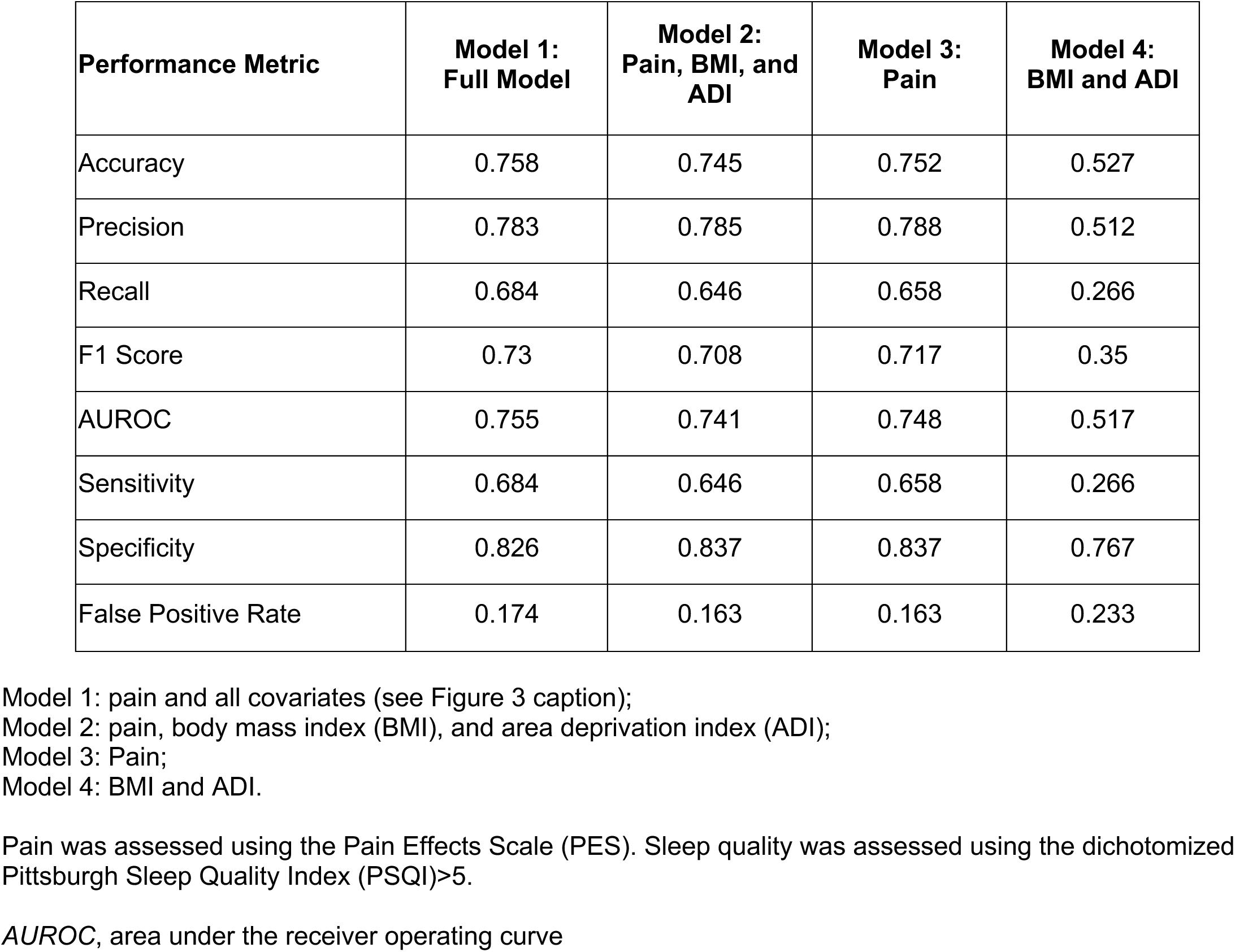
Performance of the least absolute shrinkage and selection operator (LASSO) logistic regression models predicting baseline poor sleep quality.

**S-Table 7.** Feature coefficients of the least absolute shrinkage and selection operator (LASSO) logistic regression models predicting baseline poor sleep quality.

| **LASSO coefficients** | **Model 1:**<br>**Full Model** | **Model 2:**<br>**Pain, BMI, and**<br>**ADI** | **Model 3:**<br>**Pain** | **Model 4:**<br>**BMI and ADI** |
| --- | --- | --- | --- | --- |
| Pain | 0.039 | 0.038 | 0.039 |  |
| Age | -0.004 |  |  |  |
| Gender (Male) | -0.025 |  |  |  |
| Race / Ethnicity | 0 |  |  |  |
| Disease duration | 0.002 |  |  |  |
| Body mass index | 0.003 | 0.003 |  | 0.006 |
| Elixhauser comorbidity index | 0.002 |  |  |  |
| Disease modifying therapy efficacy - Standard | 0 |  |  |  |
| Disease modifying therapy efficacy - High | -0.017 |  |  |  |
| Area deprivation index | 0 | 0 |  | 0 |

Model 1: pain and all covariates (see Figure 3 caption);
Model 2: pain, body mass index (BMI), and area deprivation index (ADI);
Model 3: Pain;
Model 4: BMI and ADI.
Pain was assessed using the Pain Effects Scale (PES). Sleep quality was assessed using the dichotomized Pittsburgh Sleep Quality Index (PSQI)>5.

**S-Table 8.**
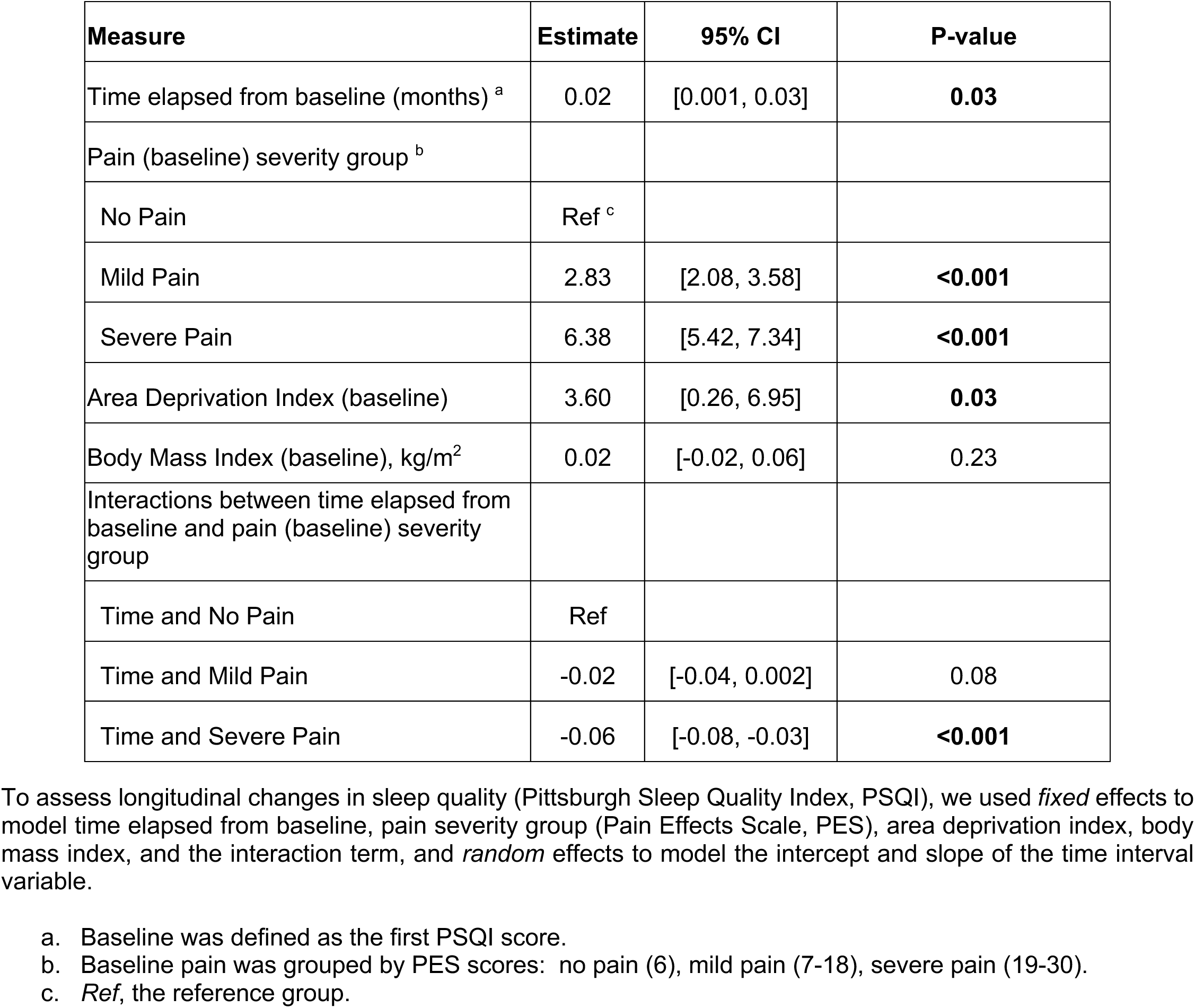
Covariate-adjusted linear mixed-effects model examining the association between baseline pain (Pain Effects Scale score categories) and longitudinal changes in sleep quality (in the longitudinal multiple sclerosis cohort).

| Measure | Estimate | 95% CI | P-value |
| --- | --- | --- | --- |
| Time elapsed from baseline (months) <sup>a</sup> | 0.02 | [0.001, 0.03] | **0.03** |
| Pain (baseline) severity group <sup>b</sup> |  |  |  |
| No Pain | Ref <sup>c</sup> |  |  |
| Mild Pain | 2.83 | [2.08, 3.58] | **<0.001** |
| Severe Pain | 6.38 | [5.42, 7.34] | **<0.001** |
| Area Deprivation Index (baseline) | 3.60 | [0.26, 6.95] | **0.03** |
| Body Mass Index (baseline), kg/m<sup>2</sup> | 0.02 | [-0.02, 0.06] | 0.23 |
| Interactions between time elapsed from baseline and pain (baseline) severity group |  |  |  |
| Time and No Pain | Ref |  |  |
| Time and Mild Pain | -0.02 | [-0.04, 0.002] | 0.08 |
| Time and Severe Pain | -0.06 | [-0.08, -0.03] | **<0.001** |

To assess longitudinal changes in sleep quality (Pittsburgh Sleep Quality Index, PSQI), we used *fixed* effects to model time elapsed from baseline, pain severity group (Pain Effects Scale, PES), area deprivation index, body mass index, and the interaction term, and *random* effects to model the intercept and slope of the time interval variable.
- a. Baseline was defined as the first PSQI score. - b. Baseline pain was grouped by PES scores: no pain (6), mild pain (7-18), severe pain (19-30). - c. *Ref*, the reference group.

**S-Table 9.**
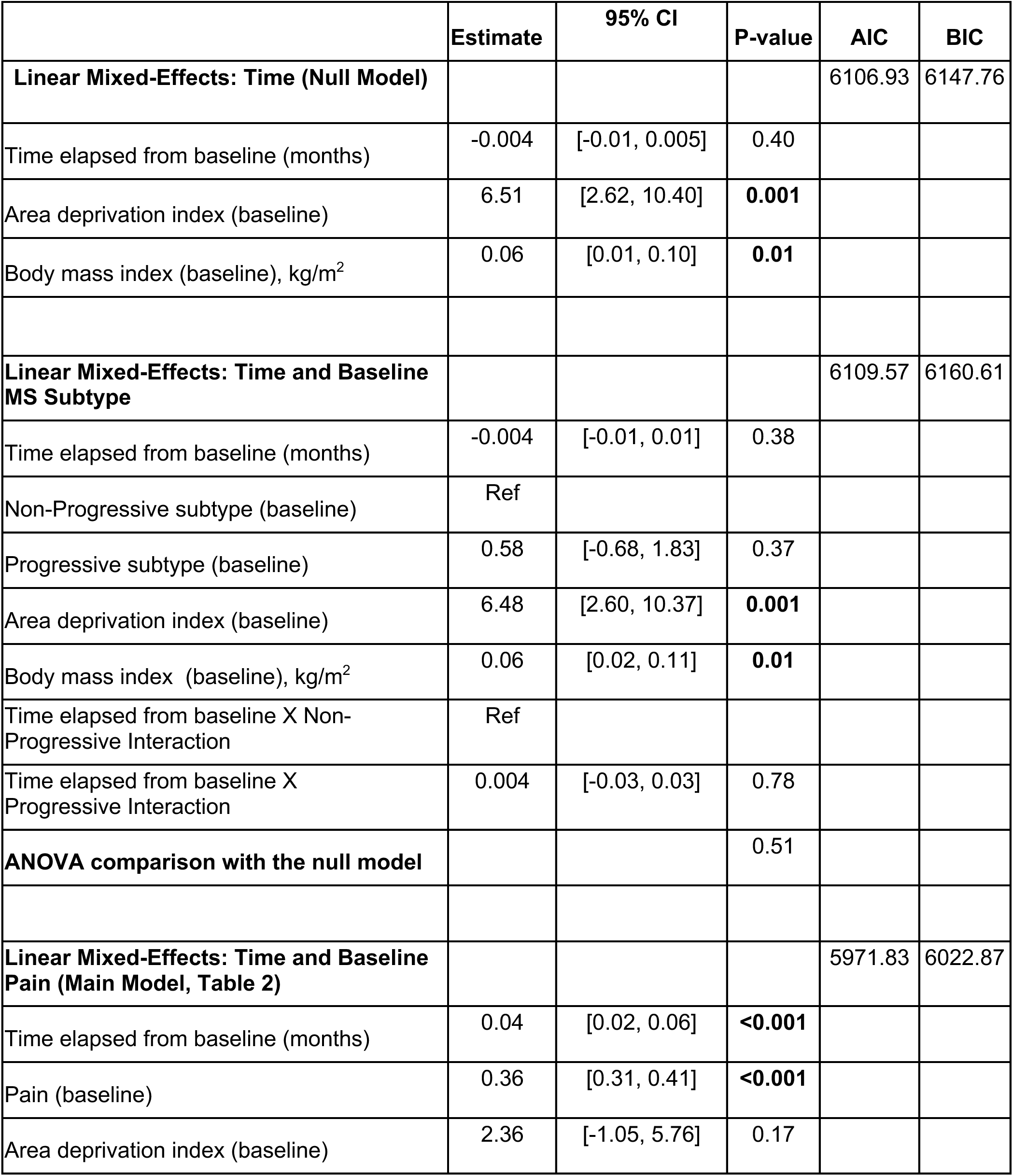
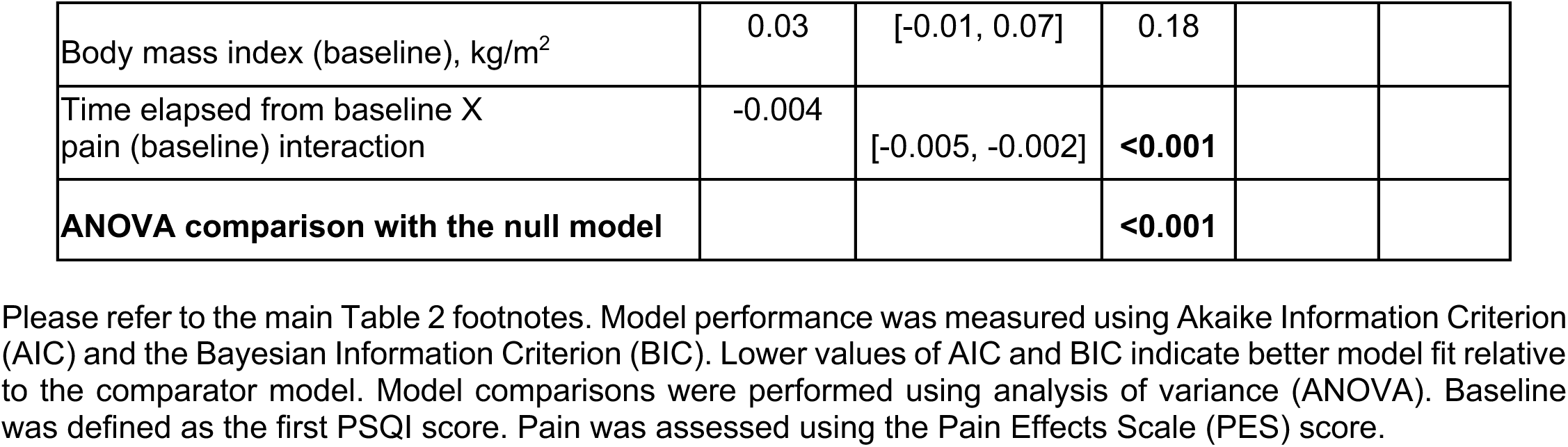
Additional covariate-adjusted linear mixed-effects models to assess longitudinal sleep quality changes (null, MS subtype, and baseline pain).

**S-Table 10.** Covariate-adjusted generalized estimating equations (GEE) to assess longitudinal sleep quality changes (null, MS subtype, and baseline pain).

|  | Estimate | 95% CI | P-value | QIC |
| --- | --- | --- | --- | --- |
| **GEE: Time (Null Model)** |  |  |  | 21755.49 |
| Time elapsed from baseline (months) | 0.002 | [-0.01, 0.01] | 0.77 |  |
| Area deprivation index (baseline) | 6.08 | [2.47, 9.69] | 0.001 |  |
| Body mass index (baseline), kg/m<sup>2</sup> | 0.06 | [0.02, 0.10] | 0.01 |  |
| **GEE: Time and Baseline MS Subtype** |  |  |  | 21698.40 |
| Time elapsed from baseline (months) | < -0.001 | [-0.01, 0.01] | 1.00 |  |
| Non-Progressive subtype (baseline) | Ref |  |  |  |
| Progressive subtype (baseline) | 0.36 | [-0.64, 1.36] | 0.48 |  |
| Area deprivation index (baseline) | 6.19 | [2.60, 9.77] | 0.001 |  |
| Body mass index (baseline), kg/m<sup>2</sup> | 0.06 | [0.02, 0.10] | 0.01 |  |
| Time elapsed from baseline X Non-Progressive Interaction | Ref |  |  |  |
| Time elapsed from baseline X Progressive Interaction | 0.020 | [-0.02, 0.05] | 0.32 |  |
| **GEE: Time and Baseline Pain** |  |  |  | 16586.00 |
| Time elapsed from baseline (months) | 0.05 | [0.03, 0.07] | <0.001 |  |
| Pain (baseline) | 0.38 | [0.33, 0.43] | <0.001 |  |
| Area deprivation index (baseline) | 1.93 | [-0.92, 4.78] | 0.19 |  |
| Body mass index (baseline), kg/m<sup>2</sup> | 0.03 | [-0.01, 0.07] | 0.21 |  |
| Time elapsed from baseline X pain (baseline) interaction | -0.004 | [-0.01, -0.002] | <0.001 |  |

To model longitudinal sleep quality using generalized estimating equations (GEE) with independent correlation structure, we included all participants with at least 1 Pittsburgh Sleep Quality Index (PSQI) score (n= 601). Model performance was measured using Quasi-Likelihood Information Criterion (QIC) with a lower value indicating
better model fit. Baseline was defined as the first PSQI score. Pain was assessed using the Pain Effects Scale (PES) score. Ref denotes the reference group.

